# Naturalistic Outcomes with fMRI-Guided and Non-fMRI-Guided Accelerated TMS for Depression

**DOI:** 10.1101/2025.09.03.25334813

**Authors:** Danielle D. DeSouza, Nathan F. Meng, Noah P. DeGaetano, Erica Nakano, Vivian Hoang, Teresa Nguyen, David S. Ling, David M. Carreon, Shan H. Siddiqi

**Author notes:** Corresponding Author: Name: Danielle D. DeSouza Institution: Acacia Clinics, Address: 877 W. Fremont Ave., Suite N-3, Sunnyvale CA, 94087. indicates co-last-authorship (David M. Carreon, Shan H. Siddiqi). Previous Presentation: **None**. **Disclosures:** D.D.D., N.F.M., N.P.D., E.N., V.H., T.N., D.S.L., and D.M.C. receive a salary from Acacia Clinics. D.D.D., N.F.M., N.P.D., E.N., T.N., D.S.L., D.M.C. have equity/stock options in Acacia Clinics. S.H.S. is an owner of intellectual property involving the use of individualized resting-state network mapping to target TMS, which was filed in 2016, has yielded under $200 in royalties, and was not employed in the present work. S.H.S. is also a scientific consultant for Magnus Medical, received investigator-initiated research funding from Neuronetics (2019) and Brainsway (2022), and received speaking fees from Brainsway (2021) and Otsuka (for PsychU.org, 2021). S.H.S. is also a former shareholder in Brainsway (publicly traded) and a current shareholder in Magnus Medical (not publicly traded). S.H.S. is also a clinical consultant for Acacia Clinics, Kaizen Brain Center, and Boston Precision Neurotherapeutics.

## Abstract

**Objective:** To estimate the effects of accelerated transcranial magnetic stimulation (aTMS) with and without functional MRI (fMRI) guidance in a naturalistic clinic setting.

**Methods:** We retrospectively analyzed data on patients who received aTMS for depression at a single subspecialty clinic between 2019 and 2025, most completing a five-day course (up to 10 treatments per day). fMRI-guided targets were selected from resting-state connectivity to the subgenual cingulate cortex. Non-fMRI-guided targeting was done using scalp-based measurement procedures. Response was defined as a Clinical Global Impression-Improvement score ≤2 within one month post-TMS. Logistic regression evaluated predictors of response and propensity score matching was used to create comparable groups of fMRI-guided and non-fMRI-guided patients while controlling for baseline heterogeneity. Predictors included age, sex, baseline severity, past electroconvulsive therapy, past TMS, previous antidepressant trials, number of sessions, comorbidities, and fMRI guidance.

**Results:** Of 195 patients receiving aTMS, 115 received fMRI guidance and 80 did not. The overall response was 72.8% (77.4% with fMRI guidance, 66.3% without). Logistic regression identified fMRI guidance as the only independent predictor of response (p=0.038). In 71 pairs matched using propensity scores, response rates were 77.5% with fMRI guidance and 62.2% without (OR=2.30, 95% CI=1.05–5.41, p=0.035), yielding a number needed to treat of 6.5 to achieve one added response with fMRI guidance.

**Conclusions:** In this large naturalistic sample, aTMS was an effective, rapid-acting antidepressant with or without fMRI guidance. fMRI-guided targeting significantly improved outcomes.

## INTRODUCTION

Accelerated transcranial magnetic stimulation (aTMS) protocols, in which two or more TMS stimulation sessions are delivered per day, have emerged as a promising therapeutic strategy for major depression (1–6). These approaches can accelerate the onset of therapeutic benefit by delivering a full treatment course in only five days. Early clinical trials reported remission rates as high as 86% after five days of aTMS in patients with highly treatment-resistant depression (7).

Some of the most successful aTMS studies have employed resting-state functional MRI (fMRI) to identify personalized stimulation targets (7–9). For depression, fMRI can be used to identify a target within the left dorsolateral prefrontal cortex (L-DLPFC) that shows the strongest negative functional connectivity with the subgenual cingulate cortex (SGC) (10). Multiple retrospective studies have shown that TMS was more effective when the stimulation site was incidentally closer to this fMRI-derived target (11–14). However, fMRI guidance requires specialized expertise, image processing software, and neuronavigation equipment, creating a logistical hurdle for most clinics. As a result, the added value of fMRI-guided targeting is a growing topic of interest, with results from a recent randomized trial forthcoming (15).

Importantly, findings from small highly controlled trials with carefully selected patient populations may not generalize to real-world clinical populations across diverse demographic profiles, clinical characteristics, and treatment protocol variations (16–18). Here, we report clinical outcomes with fMRI-guided and non-fMRI-guided accelerated rTMS in a large, naturalistic clinical cohort. We applied multivariate logistic regression and propensity score matching to account for baseline differences between the two groups. We aimed to estimate naturalistic outcomes of aTMS and to test the hypothesis that fMRI-guided accelerated rTMS would improve the odds of clinical response.

## METHODS

### Study Design

We conducted a retrospective analysis of adult patients with depression who received aTMS at a single outpatient clinic between July 2018 and April 2025. aTMS was delivered as multiple daily sessions (≥2 per day on average) using intermittent theta-burst stimulation (iTBS) protocols. Targets included the L-DLPFC, either alone or in combination with secondary targets. fMRI guidance was introduced as an option in November 2021; no patients received fMRI guidance prior to this date. Treatment was delivered using a figure-eight coil for fMRI-guided treatments, and either a figure-eight coil or H-coil for non-fMRI-guided treatments. Pearl Institutional Review Board determined this study met exemption criteria as secondary research, and informed consent was not required.

### Participants

Eligible participants were adults (≥18 years) with unipolar or bipolar depression, meeting DSM-5 criteria for a major depressive episode without psychotic features. Diagnoses were established through semi-structured clinical interviews. Patients were included if this was their first accelerated iTBS course at the clinic. Prior conventional once-daily TMS was permitted if completed at least two weeks before the current course. Exclusion criteria included TMS or MRI contraindications and comorbid psychiatric conditions deemed more symptomatic than the depressive episode. Patients with primary anxiety-related disorders (including obsessive-compulsive disorder and posttraumatic stress disorder) were generally treated with dorsomedial prefrontal cortex (19,20) targeting and were excluded from this analysis.

### Target Generation

For patients who had resting-state fMRI-guided targets, different scanners were used depending on the patient’s geographic preference. To reconcile this discrepancy, minimum quality standards were applied, including at least 20 minutes in scan duration, 3T magnetic field, at least 45 mT/m magnetic gradient, up to 3 second repetition time, and up to 3.5 x 3.5 x 4 mm spatial resolution. Incidentally, all scans were conducted on a Siemens MAGNETOM MRI scanner (Verio, Trio, Skyra, or Prisma), although this was not an *a priori* quality criterion.

High-resolution T1-weighted anatomical scans were also acquired. Prior to resting-state connectivity analysis, scans were preprocessed as described previously (21). Steps included spatial alignment, atlas registration, ART-based motion censoring (framewise displacement threshold of 0.5 mm), global signal regression, temporal bandpass filtering (0.009 to 0.08 Hz), and spatial smoothing (6mm FWHM kernel), using SPM12 and the CONN toolbox (22). Voxel-wise (Fig. 1a) resting-state functional connectivity (rsFC) was computed from the SGC (Fig. 1b) as defined by the default mask in CONN for specific regions of interest. The resulting connectivity map (Fig. 1c) was reviewed manually by a neuropsychiatrist with expertise in MRI-guided TMS (S.H.S.). Targets were chosen based on several factors: magnitude of negative connectivity to SGC, size/shape of the target cluster relative to a TMS coil, anticipated tolerability of coil position, accessibility with TMS, and other case-by-case factors (Fig. 1c). TMS was then applied to this optimized target (Fig. 1d) using the Localite TMS Navigator system.

**Figure 1:**
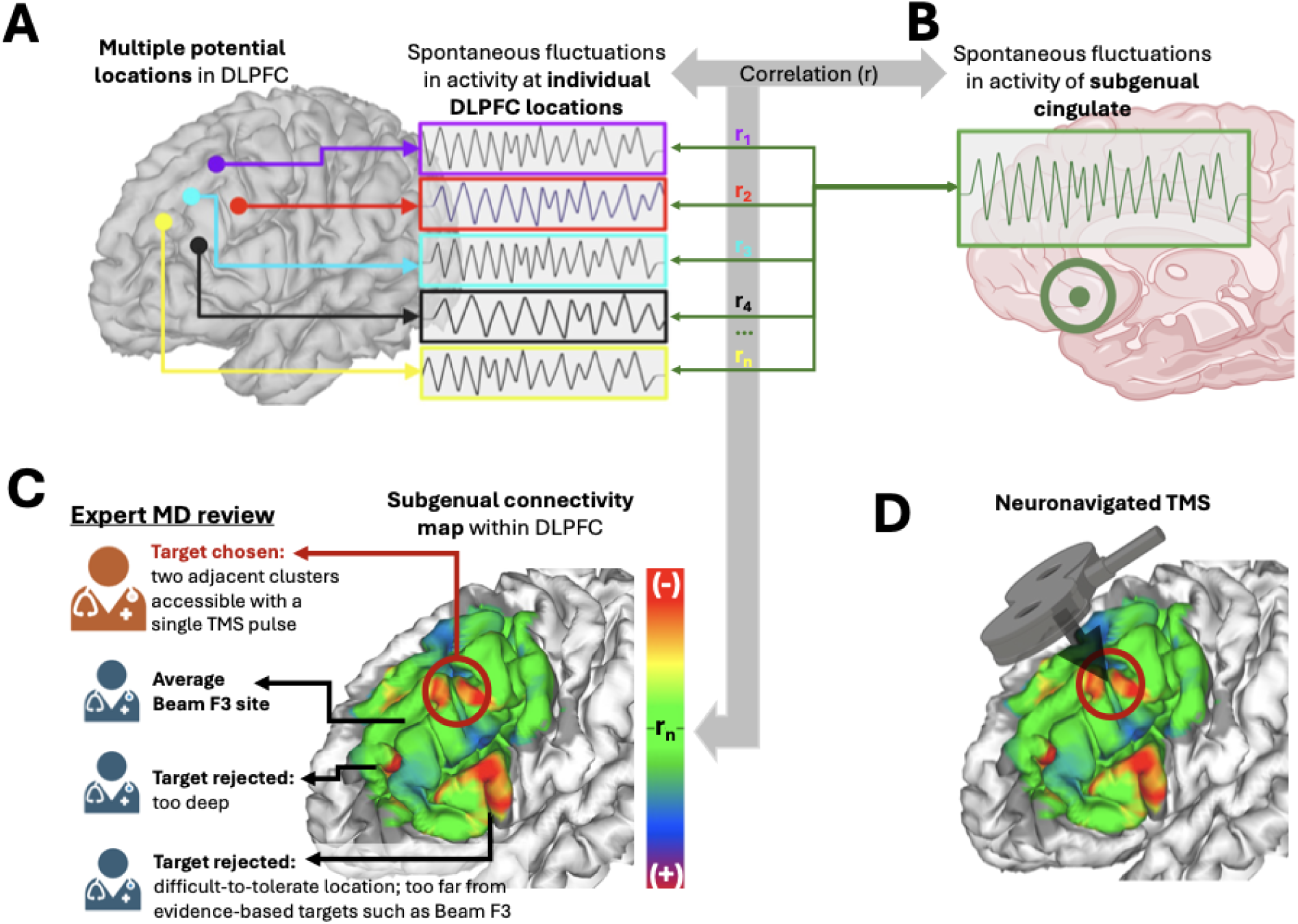
fMRI guidance procedure. **(A)** fMRI was used to measure spontaneous fluctuations in thousands of individual brain voxels in the DLPFC; five examples are depicted here. **(B)** These fluctuations were compared with the subgenual cingulate using Pearson correlations, yielding a map **(C)** of subgenual connectivity across the DLPFC. These maps were reviewed by an expert clinician to identify a TMS target **(D)**.

In patients who did not use fMRI-guided targeting, the L-DLPFC stimulation site was identified using the Beam F3 method (23), which estimates the F3 location of the 10-20 system for standardized placement of electroencephalogram (EEG) electrodes (Fig. 1c). This approach accounts for individual variability in head size but does not require neuroimaging guidance for target localization. For a subset of patients, an H-coil was used. Patients were typically allocated to Beam F3 or H-coil based on schedule availability, not on clinical factors.

In both groups, some patients also received stimulation to another target in addition to L-DLPFC, when deemed clinically appropriate.

### Clinical Assessments

The clinical global impressions improvement (CGI-I) scale was completed by board-certified clinicians at baseline, immediately post-TMS, 1 week post-TMS, and 1 month post-TMS. Evaluations were conducted in-person whenever feasible. Remission, response, and non-response were defined *a priori* as CGI-I scores of 1, ≤2, and ≥ 3 (24), respectively. The primary outcome was response or remission at one or more time points after treatment, following the clinical trial that led to FDA clearance of aiTBS (9). Baseline self-reported measures included the Patient Health Questionnaire-9 (PHQ-9) and the Generalized Anxiety Disorder 7-item scale (GAD-7).

Adverse events (AEs) were systematically monitored throughout treatment and follow-up. Patients were asked to fill out self-report surveys at the beginning of each treatment day and after each stimulation session during treatment courses. Post-treatment AEs were documented during clinical follow-up visits. Proportions of AEs experienced in various clinical categories (see Supplemental Table S1 for more details) were calculated for each group.

### Predictors of Treatment Response

We completed a binary logistic regression analysis to examine baseline predictors of response. Candidate predictors included: age, sex at birth, baseline PHQ-9 and GAD-7 scores, bipolar disorder diagnosis, fMRI-guidance, past electroconvulsive therapy (ECT), past TMS, number of previous antidepressant trials, mean number of TMS treatment sessions per day, and diagnosis group. This latter variable coded individuals with MDD only (no comorbidities), allowing us to examine the MDD-only group to emulate clinical trials that typically exclude the other categories. Continuous variables were modeled in their original units, and categorical variables as factors with prespecified level coding.

As the non-fMRI-guided group included an additional variable, TMS coil type, we did a subgroup analysis excluding individuals who had aTMS using an H-coil to remove this potential confound. Prior to model fitting, we applied a complete-case filter, excluding participants with missing values in the outcome or any candidate predictors. To address multicollinearity, we used an iterative variance inflation factor (VIF) pruning procedure. At each iteration, we calculated VIFs from a dummy-coded design matrix to remove any predictors with VIF > 5. The final model included the treatment indicator (fMRI-guided vs. non-fMRI-guided) and all retained predictors after pruning. The model was fit using a logistic regression with a binomial family and logit link, and odds ratios (ORs) with 95% Wald confidence intervals were computed for all predictors.

### Comparative Effectiveness of fMRI Guidance Using Matched Analysis

A propensity score analysis was used to compare outcomes between the fMRI-guided and non-fMRI-guided groups while matching cases (fMRI guidance) against controls (scalp-based targeting) based on measured factors. This approach created matched pairs across the two groups, emulating the balance of a randomized study design (25). The propensity score was defined as the probability of receiving fMRI-guided treatment using the same covariates included in the logistic regression model. Propensity scores were estimated using a logistic regression model (binomial family, logit link) with maximum likelihood estimation.

Matching was performed using nearest neighbor matching with replacement, applying a caliper width of 0.20 on the logit of the propensity score (25,26), and restricting any participant to be matched no more than twice (27). We additionally conducted sensitivity analyses testing caliper widths of 0.25, 0.30, 0.40, and 0.50, to examine the stability of matched samples and estimated treatment effects across a range of caliper specifications. Covariate balance after matching was assessed using standardized mean differences (SMDs) for each covariate. An SMD <0.10 was considered adequate balance between the fMRI-guided and non–fMRI-guided groups, whereas SMD values >0.20 were considered evidence of serious imbalance (25,28). Between-group differences in the primary outcome in the matched sample were compared using McNemar’s exact test for paired binary data, and odds ratios with 95% confidence intervals were calculated.

As in the logistic regression analysis, we performed an exploratory subgroup propensity score analysis limited to patients who received aTMS with a figure-eight coil, excluding those in the non-fMRI-guided group who had been treated with an H-coil to remove this potential confound. All analyses were conducted in R (version 4.5.1).

## RESULTS

### Participant Characteristics

A total of 246 participants met inclusion and exclusion criteria (135 fMRI-guided, 111 non–fMRI-guided). For hypothesis testing, analyses were restricted to the 195 participants (115 fMRI-guided, 80 non-fMRI-guided) with complete data on all variables of interest, to ensure robust and unbiased estimation (Table 1). These participants were included in both the logistic regression and propensity score analyses.

**Table 1:**
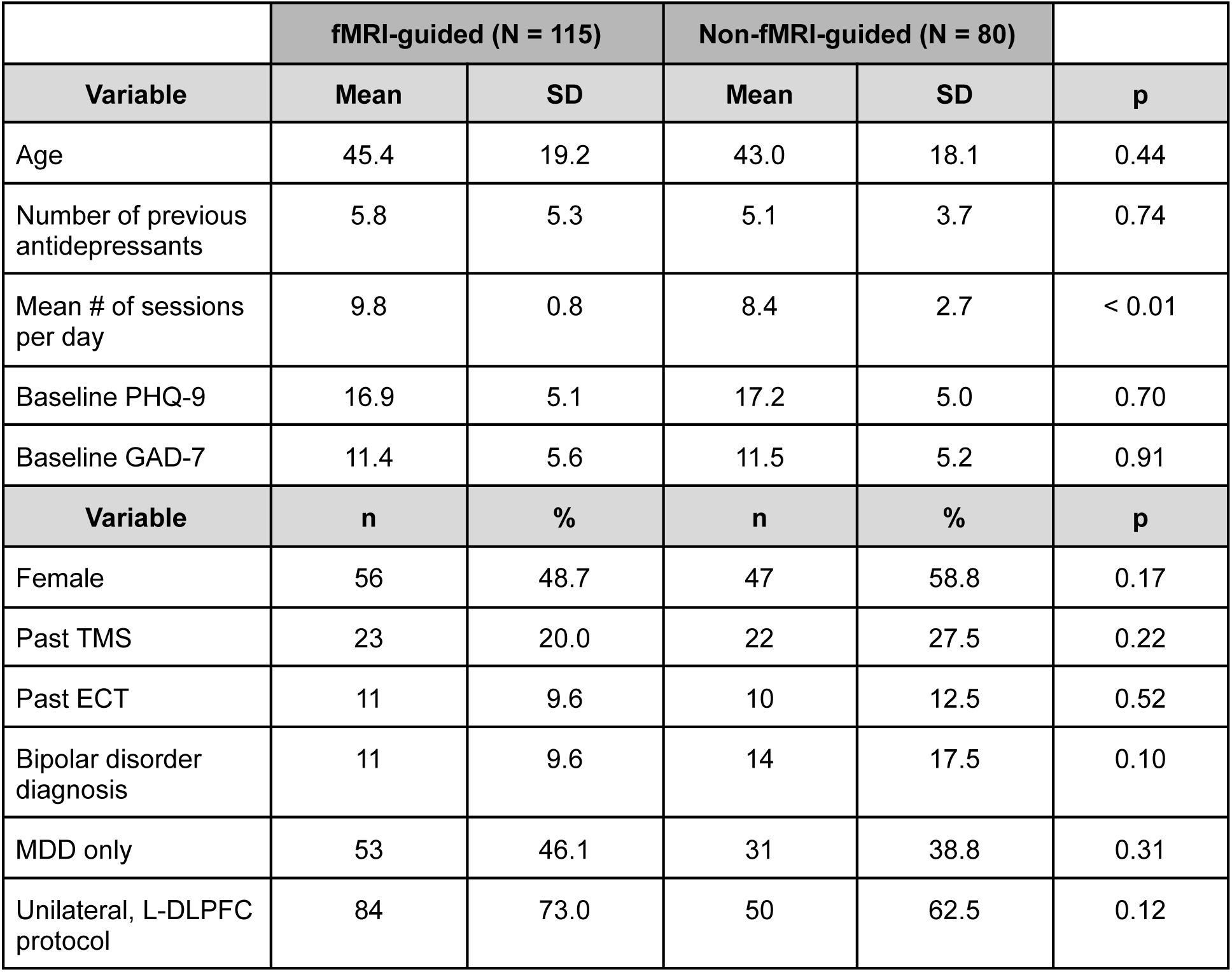
Patient Characteristics in the fMRI-guided and Non-fMRI-guided groups Baseline demographic and clinical characteristics of patients receiving fMRI-guided and non-fMRI-guided rTMS. Continuous variables are presented as mean ± SD; categorical variables as patient count (n) and proportion (%). Analyses were performed on the full sample unless otherwise indicated in parentheses, reflecting reduced sample sizes due to missing data. For two patients, treatment was provided for MDD but comorbidity data were unavailable. Between-group comparisons used Welch’s t-test or Wilcoxon rank-sum tests for continuous variables and χ² or Fisher’s exact tests for categorical variables.

### Treatment Outcomes

The overall response rate (CGI-I ≤ 2) was 72.8%. Response was higher in the fMRI-guided group (77.4%, n=115) than the non-fMRI-guided group (66.3%, n=80) (Fig. 2a); however this difference was not statistically significant (Fisher’s exact test: 95% CI=0.92–3.30, p=0.10). More than half of all patients (51.8%) achieved a CGI-I score of 1 (very much improved). Within the non-fMRI-guided group, response rates were 69.1% among patients treated with a figure-eight coil (n=55) and 60.0% among those treated with an H-coil (n=25). Of those treated with an H-coil, the majority (n=22, 88%) were treated with an H1-coil, while the remainder received treatment using an H7-coil tilted to stimulate the L-DLPFC.

**Figure 2:**
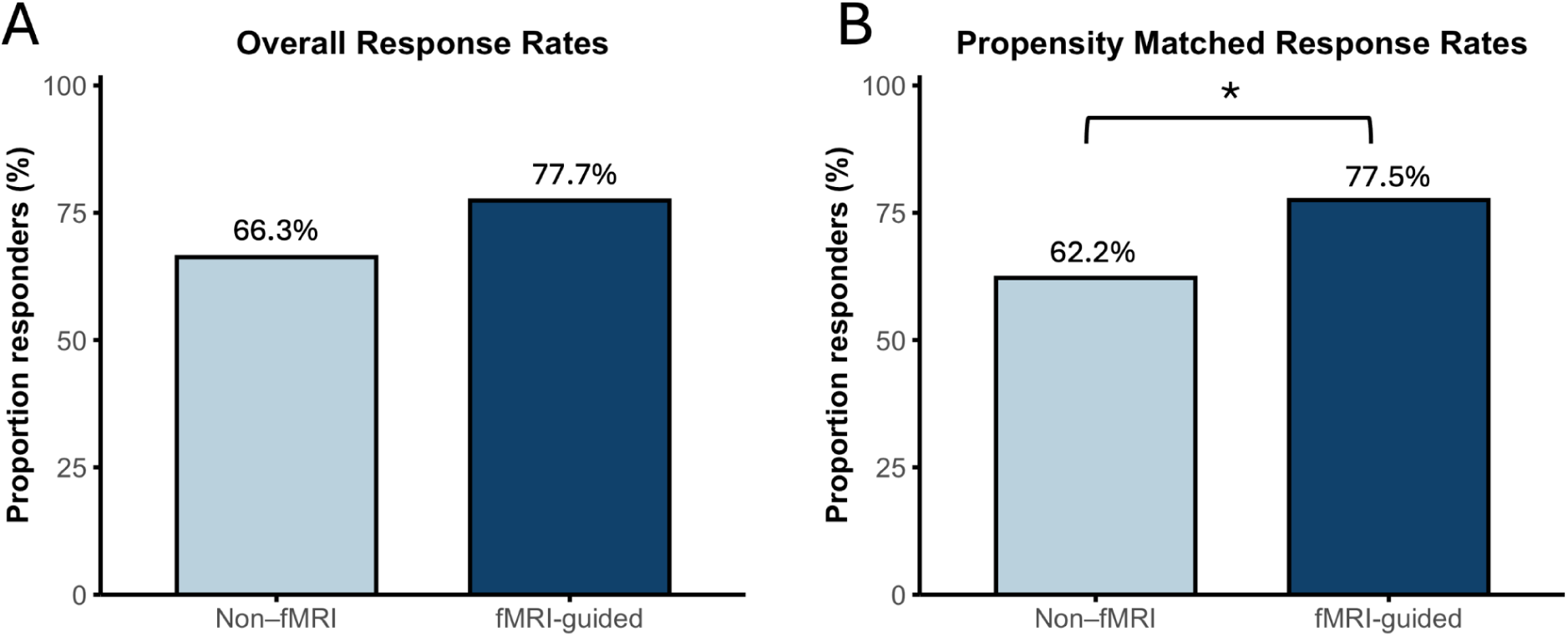
Accelerated TMS Treatment Outcomes by Targeting Approach. **(A)** Overall response rates were higher in the fMRI-guided aTMS group compared with the non-fMRI-guided group, but this difference was not statistically significant. **(B)** After balancing baseline characteristics using propensity score matching, fMRI-guided patients demonstrated significantly greater likelihood of achieving response (OR=2.3). Matched response rates shown for unique patient proportions. Asterisk indicates p<0.05.

### Predictors of Clinical Outcome

In the logistic regression analysis, fMRI-guided targeting was associated with greater odds of achieving response on the CGI-I compared with non-fMRI-guided treatment (OR = 2.13, 95% CI = 1.05–4.34, p=0.038). Greater number of previous antidepressant trials showed a trend toward lower odds of response (OR=0.93, 95% CI=0.87–1.00, p=0.065). No other baseline variables were significantly associated with outcome (all p>0.05) (Fig. 3).

**Figure 3:**
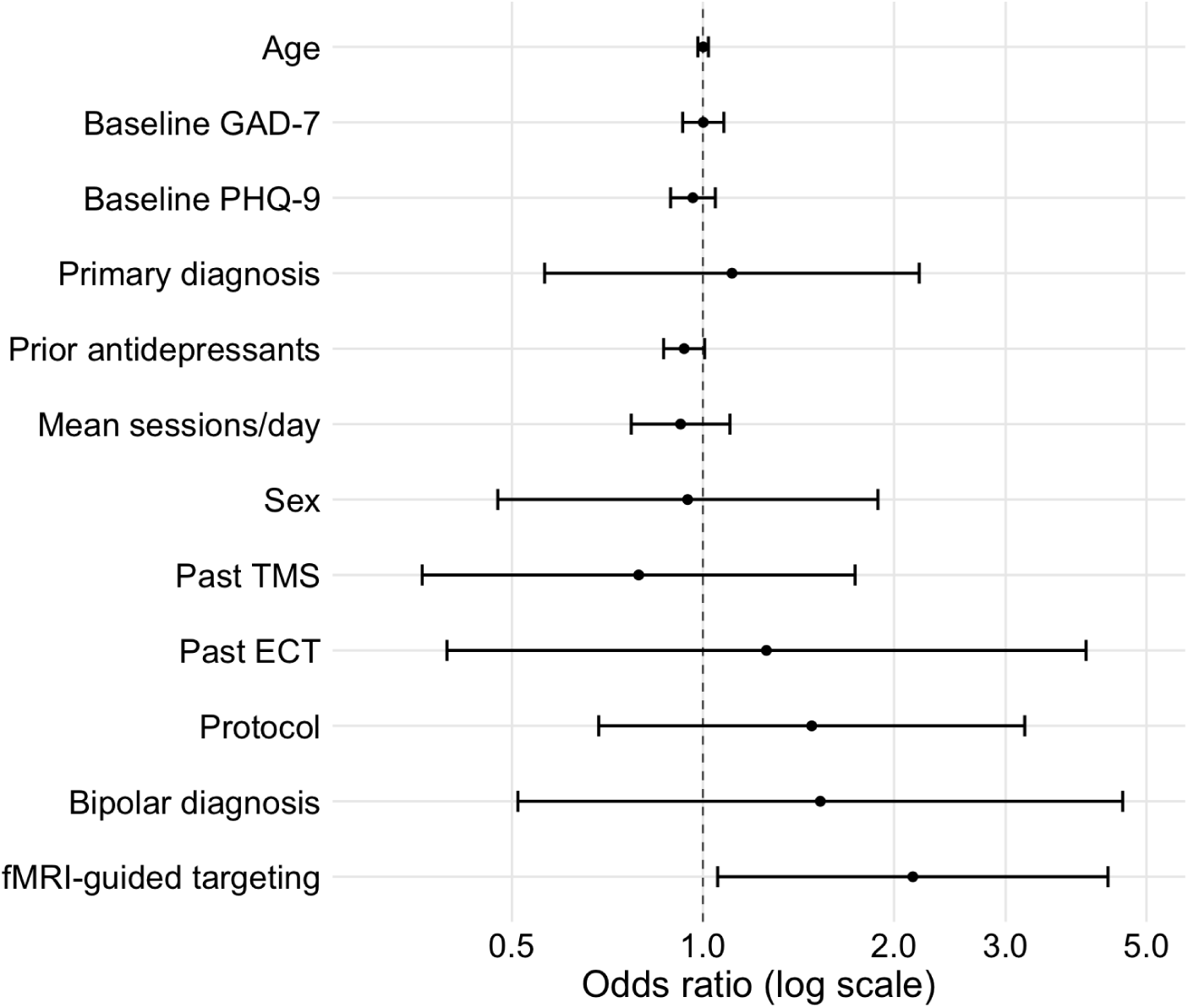
Adjusted Odds Ratios for Predictors of Treatment Response. Forest plot showing adjusted odds ratios (ORs) and 95% confidence intervals for each predictor from the multivariable logistic regression model in the full sample (n= 195). Confidence intervals crossing OR=1 suggest no statistically significant association.

In a subgroup analysis limited to patients treated with a figure-eight coil (n = 170), the number of previous antidepressant trials was significantly associated with outcome, with lower odds of response observed for patients with more prior trials (OR=0.91, 95% CI=0.84–0.99, p=0.029). No other baseline variables were significant (all p>0.05).

### Propensity Score Results

All covariates met the <0.10 balance criterion, with post-match SMDs ≤0.09, except for baseline GAD-7 score. The GAD-7 SMD was 0.13, slightly above the threshold but well below the >0.20 benchmark for serious imbalance, indicating that matching effectively minimized baseline differences between groups.

For the full-sample analysis (n=195), propensity score matching (caliper = 0.20, reuse limited to 2) yielded 71 matched pairs. On the pair scale (n=142 observations; 71 fMRI-guided, 71 non-fMRI-guided), the response rate was 77.5% (55/71) with fMRI guidance and 53.5% (38/71) without. The proportion in the non-fMRI-guided group reflects the reuse of some patients across pairs (due to smaller sample size in this group), with reused controls more often being non-responders. When calculated on a unique-patient basis, ensuring that non-responders were not overrepresented in the non-fMRI group, the corresponding proportions were 77.5% (55/71) for the fMRI-guided group and 62.2% (28/45) for the non-fMRI-guided group (Fig. 2b), representing an absolute risk reduction of 15.3% and a number needed to treat of 6.6 to achieve one response. McNemar’s exact test indicated that the odds of response were 2.3 times higher with fMRI-guided targeting (95% CI, 1.05–5.41; p=0.035). In sensitivity analyses, widening the caliper up to 0.50 yielded identical matched samples (71 pairs), covariate balance (maximum SMD=0.13), and effect estimates (OR=2.30; 95% CI 1.05–5.41), confirming the robustness of the primary findings.

In the subgroup treated with a figure-eight coil (n=170), all covariates met the <0.10 balance criterion, with post-match SMDs ≤ 0.09 except for the number of previous antidepressants (SMD =0.10). Matching with the same parameters (caliper = 0.20, reuse limited to 2) yielded 56 matched pairs. On the pair scale (n=112 observations; 56 fMRI-guided, 56 non-fMRI-guided), 76.8% (43/56) of the fMRI guided group and 46.4% of the non-fMRI-guided group (26/56) achieved a response. When calculated for unique patients, the corresponding proportions were 83.9% (47/56) for the fMRI-guided group and 66.7% (22/33) for the non-fMRI-guided group, representing an absolute risk reduction of 17.2% and a number needed to treat of 5.8 to achieve a response. McNemar’s exact test indicated that the odds of response were 3.4 times higher with fMRI-guided targeting (95% CI, 1.20–11.79; p=0.017).

### Safety

Treatment was well-tolerated with no adverse event (AE)-related discontinuations (Table 2). The most common AEs were headache, fatigue, and treatment-emergent anxiety. Having any AE was more common in the fMRI-guided group (p=0.016), as were headache (p=0.026), cognitive effects (p=0.046), and stimulation site pain (p=0.046). In secondary analysis, somatic symptoms and treatment-emergent anxiety were combined, consistent with prior work suggesting that these symptoms may be exacerbated by the anti-SGC target (19). This combined category was more common in the fMRI-guided group (24.3% vs 12.2%, OR 2.32 [1.02–5.28] p=0.041). In all but two cases, treatment-emergent anxiety self-resolved before the end of treatment week. The remaining two patients improved within one month after right Beam TMS to right Beam and therapy. Two older participants experienced falls: an individual in their 70s with a prior history of falls fell during treatment week, and an individual in their 90s fell two days post-treatment. Neither reported dizziness or unsteadiness during stimulation, and neither were directly attributable to TMS. One participant tragically completed suicide two days post-treatment. This individual had been recognized as very high-risk at baseline, with two prior serious suicide gestures. A detailed case review by three board-certified psychiatrists, determined it unrelated to direct TMS effects.

**Table 2:** Adverse Events by Treatment Group Adverse events (AEs) among patients who received fMRI-guided and non-fMRI-guided aTMS. Group proportions were compared with a two-sided pooled two-proportion z-test. * p<0.05.

| Outcome | fMRI-Guided (%)<br>(N = 111) | Non-fMRI-guided (%)<br>(N = 74) | Risk difference (%)<br>[95% CI] | p |
| --- | --- | --- | --- | --- |
| Any adverse event | 78.4 | 62.2 | 16.2 [2.8, 29.7] | 0.016* |
| Headache | 61.3 | 44.6 | 16.7 [2.2, 31.2] | 0.026* |
| Fatigue | 31.5 | 29.7 | 1.8 [-11.7, 15.3] | 0.795 |
| Anxiety | 15.3 | 6.8 | 8.6 [-0.3, 17.4] | 0.078 |
| Gastrointestinal | 14.4 | 12.2 | 2.3 [-7.7, 12.2] | 0.661 |
| Somatic Sensations | 13.5 | 5.4 | 8.1 [-0.1, 16.3] | 0.075 |
| Dizziness /<br>Lightheaded | 9.9 | 5.4 | 4.5 [-3.1, 12.1] | 0.272 |
| Mood Dip | 9.0 | 9.5 | -0.5 [-9.0, 8.1] | 0.917 |
| Cognitive | 8.1 | 1.4 | 6.8 [1.0, 12.5] | 0.046* |
| Sensory Change | 8.1 | 5.4 | 2.7 [-4.5, 9.9] | 0.481 |
| Site Pain | 8.1 | 1.4 | 6.8 [1.0, 12.5] | 0.046* |
| Twitching | 6.3 | 5.4 | 0.9 [-6.0, 7.8] | 0.800 |
| Irritability / Agitation | 5.4 | 8.1 | -2.7 [-10.2, 4.8] | 0.465 |
| Tinnitus | 4.5 | 4.1 | 0.5 [-5.5, 6.4] | 0.883 |
| Sleep Change | 3.6 | 4.1 | -0.5 [-6.1, 5.2] | 0.875 |
| Jaw Pain | 1.8 | 2.7 | -0.9 [-5.3, 3.5] | 0.680 |

## DISCUSSION

Since the recent emergence of aTMS as a rapid-acting antidepressant treatment, there have been growing questions about its utility in a naturalistic sample of patients with multiple comorbidities. In this large, single-site observational cohort of patients, aTMS produced a 72.8% response rate within one month despite a high degree of baseline treatment resistance (mean >5 medication failures). Additionally, more than half of patients experienced remission of symptoms. After propensity matching, response rates were high with and without fMRI guidance, although fMRI guidance led to significantly better outcomes. These findings support the view of aTMS as a highly effective rapid-acting treatment for patients with treatment-resistant major depression.

These response rates are modestly lower than rates previously reported with fMRI-guided aTMS (29,30). There are several possible explanations for this observation. First, our results are consistent with the well-known tendency of large naturalistic studies to report smaller effects than early clinical trials (31). Second, our sample included patients with multiple comorbidities, and often the primary diagnosis was less clearly defined. As a result, many of these patients would have been excluded from prospective clinical trials. Third, our fMRI guidance algorithm was not identical to the one used in prior clinical trials, although it was similar to approaches associated with better antidepressant efficacy reported in retrospective studies (12,14).

The value of fMRI guidance for TMS targeting has been debated. Some retrospective studies have shown that treatment outcomes are better when stimulation sites are incidentally closer to a retrospectively-derived fMRI-guided target (32). Other work has demonstrated only modest effect sizes of personalization when based on relatively brief scans on older scanners with lower signal-to-noise in the SGC (33). These findings highlight the value of high-quality fMRI acquisition and analysis. Here, we observed that the odds of clinical response or remission were significantly greater in patients with personalized fMRI-based targeting than similar patients without fMRI-based targeting. However, response rates were high in both groups, suggesting that aTMS may still be a valuable treatment when MRI guidance is unavailable.

This result should be interpreted with caution given baseline group differences. fMRI-guided targeting was introduced midway through data collection, raising the possibility of chronological bias from unobserved time trends (34). Early aTMS adopters may have been more susceptible to placebo effects and fMRI guidance itself could also contribute to placebo response. Still, the intensity, cost, and novelty of aTMS may already approach the upper limit of placebo response regardless of fMRI. Other factors such as treatment resistance, personality traits, or financial constraints may have impacted treatment decisions or outcomes. This concern is partly mitigated by the consistent fMRI advantage across both analytic approaches: logistic regression offered greater power through covariate adjustment, whereas propensity score matching emulated certain features of randomization. Nevertheless, unmeasured confounders may remain, and head-to-head randomized trials are required to confirm these findings.

Importantly, we used fMRI to optimize targets, but not to identify biomarkers that predict or track treatment response. This decision was made for several reasons. First, localization of specific sites with fMRI (requisite for targeting) is more reliable than measuring the magnitude of functional connectivity (requisite for biomarkers). Second, in a treatment-seeking patient, treatment targets are modifiable based on fMRI localization, while the role of biomarkers in treatment decision-making is less clear. Third, SGC connectivity has been replicated in multiple clinical trials to predict efficacy of stimulation sites, while different biomarker studies yield different candidate predictors (35,36). Finally, fMRI-guided targeting was employed in the aTMS clinical trial that led to FDA clearance, while biomarkers were not. Future studies in larger samples may reveal useful biomarkers.

Treatment was generally well-tolerated, with no AE-related discontinuations (37). Transient treatment-emergent anxiety and somatic side effects were significantly elevated in the fMRI-guided group and were higher than we expected in absolute terms. In the present analysis, patients with more severe anxiety systems were excluded, making this finding even more surprising. Similar rates have been reported with neuronavigated TMS to similar targets, and anxiety exacerbations have been linked to incidental stimulation of a circuit that largely overlaps with the target employed in the present study (38). Headaches and site pain were also more common in the fMRI-guided group, although target-dependent headaches have not previously been reported to our knowledge. These findings raise the possibility that greater precision in stimulation may be associated with both enhanced efficacy and a higher incidence of certain AEs. This also mitigates concern regarding placebo effects with fMRI guidance, as there was no expectancy of differences in adverse effects.

One patient unfortunately died by suicide within one month of neuronavigated treatment. This patient was identified as high-risk before treatment, including a history of two serious suicide gestures. After extensive review of the case by three board-certified psychiatrists (D.M.C., N.P.D., S.H.S.), this death was determined to be likely unrelated to treatment. However, this case highlights the importance of ongoing suicide risk monitoring after failure of an intensive treatment that may have been perceived as a last resort.

Study strengths include the large, naturalistic sample, application of two complementary statistical approaches to address baseline heterogeneity, and consistent effect estimates across methods and sensitivity analyses. Perhaps the most important limitation in the analysis of fMRI effects, as discussed above, is the potential for unmeasured confounders, which can only be addressed by a prospective clinical trial. The fMRI analysis is also limited by the fact that all participants were stimulated at the anti-SGC target, but different targets may benefit patients with distinct comorbidities (19,39). Precision of effect sizes is also limited by the single-site design and reliance on observational data with subjective clinician-report scales. Generalizability is constrained by data collection prior to third-party payer reimbursement for aTMS, resulting in a cohort that disproportionately included individuals with greater financial resources. These limitations may be addressed by prospectively enrolling patients in a pre-registered externally-funded multi-site study with a detailed phenotyping and multiple stimulation targets. This study is now underway (NCT06376734) (40).

Overall, these results suggest that aTMS is a highly effective intervention for patients with treatment-resistant major depression. fMRI guidance led to improved rates of clinical response and transient adverse effects. In combination with randomized clinical trials, these data may support more widespread adoption of aTMS with or without fMRI guidance.

## Data Availability

All data produced in the present study are available upon reasonable request to the authors.

## Acknowledgements

We are thankful to the patients whose clinical data were included in this study. We also acknowledge discussions to conceptualize this project in collaboration with Nolan Williams, Brandon Bentzley, Owen Muir, and Michael Fox. We appreciate the contributions of the dedicated technicians, administrative, and other staff at Acacia Clinics, whose day-to-day efforts in patient care and data collection made this work possible. In addition, the investigators were funded by the following sources: S.H.S.: the NIH (PI on grants K23MH121657 and R01MH136248, co-I on grant R01MH113929), BrainsWay (Investigator-initiated grant), and Brigham Ignite.

