## Supplemental Table 1 for "Naturalistic Outcomes with fMRI-Guided and Non-fMRI-Guided Accelerated TMS for Depression"

**Table S1: Examples of Patient-Reported Adverse Event Responses by Clinical Category**

| Adverse Event Category | Example Response |
|---|---|
| **Headache** | “headache,” “headache and soreness,” “migraine” |
| **Fatigue** | “very tired,” “fatigue” |
| **Anxiety** | “panic attack,” “anxious,” “anxiety, stress, meltdown” |
| **Gastrointestinal** | “very slight stomach queasiness,” “intestinal cramping,” “nausea,” “hungry” |
| **Somatic Sensations** | “muscle cramps,” “being hot,” “I had to use my inhaler,” “peace, like a river… all of my back,” “slight burning sensation in my forehead,” |
| **Dizziness / Lightheaded** | “feeling a little bit woozy,” “slightly light-headed,” occasional balance issues” |
| **Mood Dip** | “very depressed, hard crying,” “felt down after treatment yesterday” |
| **Cognitive** | “moderate memory problems,” “forgetful,” “great increase in 'brain fog’” |
| **Sensory Change** | “colors seem more vibrant,” “my senses are somewhat heightened,” “sensitive ears,” “vision seemed unusually sharp” “increased sensitivity to sounds and light” |
| **Site pain** | “reluctant anticipation of next pulse,” “sore head,” “area that's being stimulated kinda hurts more with each session but not terribly” |
| **Twitching** | “twitching muscles (especially during TMS rounds),” “twitchiness,” “jaw twitching” |
| **Irritability / Agitation** | “very irritable,” “mild frustration,” “more agitation/irritability than normal” |
| **Tinnitus** | “slight ringing in ears,” “slight tinnitus,” “ringing in right ear for ~5min” |
| **Sleep Change** | “insomnia,” “inability to stay asleep,” “extreme sleepiness” |
| **Jaw Pain** | “jaw just a little sore,” “jaw tension” |
